# COVID-19 UK Lockdown Forecasts and R_0_

**DOI:** 10.1101/2020.04.07.20052340

**Authors:** Greg Dropkin

## Abstract

**Introduction:** The first reported UK case of COVID-19 occurred on 31 January 2020, and a lockdown of unknown duration began on 24 March. One model to forecast disease spread depends on clinical parameters and transmission rates. Output includes the basic reproduction number **R**_**0**_ and the log growth rate **r** in the exponential phase.

**Methods:** UK data on reported deaths is used to estimate **r**. A likelihood for the transmission parameters is defined from a gaussian density for **r** using the mean and standard error of the estimate. Parameter samples from the Metropolis-Hastings algorithm lead to an estimate and credible interval for **R**_**0**_ and forecasts for severe and critical cases, and deaths during a lockdown.

**Results:** In the exponential phase, the UK growth rate for log(deaths) is **r** = 0.224 with s.e. 0.005. **R**_**0**_ = 5.81 with 90% CI (5.08, 6.98). In a 12 week lockdown from 24 March with transmission parameters reduced to 20% of their previous values, around 63,000 severely ill patients will need hospitalisation by mid June, 37,000 critically ill will need intensive care, whilst over 81,000 are expected to die. Had the lockdown begun on 17 March around 16,500 severe, 9,250 critical cases and 18,500 deaths would be expected by early June. With 10% transmission, severe and critical cases peak in April.

**Discussion:** The **R**_**0**_ estimate is around twice the value quoted by the UK government. The NHS faces extreme pressures, even if transmission is reduced ten-fold. An earlier lockdown could have saved many lives.

## Introduction

Although the first two confirmed UK cases of the novel coronavirus disease COVID-19 were identified on 31 January 2020, the government hesitated for some time whilst reported cases grew to 6,650, before instituting a lockdown on 24 March. The lockdown remains in place, with no declared end date.

In describing the spread of an infectious disease, one key parameter is the basic reproduction number **R**_**0**_, the expected number of individuals who will be infected by a single infectious person if the rest of the population is susceptible. If **R**_**0**_ > 1, the disease spreads, whilst if **R**_**0**_ < 1 it will die out. Recent preprints [1, 2, 3] and published articles [4, 5] have estimated **R**_**0**_ for COVID-19, but there is no clear consensus, and the true value may depend on social characteristics of the population.

As the disease progresses, patients who recover may acquire immunity, lessening the pool of susceptible individuals. However, at the initial stages almost everyone remains susceptible, and case numbers and deaths grow exponentially. A second key descriptive parameter is **r**, the rate of increase in log(cases) or log(deaths) during this exponential growth phase.

The disease spread can be approximated with a deterministic compartmental SEIR model [6] based on the numbers of patients who are Susceptible, Infected, Infectious (mild), Infectious (severe), Infectious (critical), Recovered, or Dead, and the rates of transition between these states. As a set of linked differential equations, the SEIR model can be integrated by numerical methods to forecast the future course of the disease.

Some of the transition rates in the model can be regarded as clinical parameters. For example, the rate at which people move from infected to infectious depends on the average length of the pre-infectious period, which is a clinical characteristic and might be similar in all populations. By contrast, the rate at which mildly ill people infect others depends on the pattern of social interaction in the community. These transmission parameters are likely to vary between countries.

The UK data underestimates the number of cases, as there has been no testing in the community, and the public has been told not to inform the NHS if they feel only mildly ill. It seems likely that reported COVID-19 deaths in hospital are a more accurate reflection of mortality, though they will still be underestimates as they exclude deaths in care homes.

The approach in this paper begins by using the early data on deaths to estimate **r** directly. The default clinical parameters in the SEIR model are then taken as fixed, and posterior samples of the transmission parameters are obtained, making use of the mean and standard error of the estimate for **r** and the fact that **r** is also an outcome of the model, conditional on the parameters. Using the model, these samples also generate samples for **R**_**0**_, giving an estimate and credible interval. Finally, the samples can be applied to forecast the spread of the disease in the context of a lockdown of specified effectiveness and duration.

## Materials and methods

A time series of reported COVID-19 cases and deaths in the UK is available from Public Health England. [7] Data on log(deaths) through to 31 March was examined for linearity, and a linear model M1 was then fitted to the appropriate portion, as shown in Figure 1.

**Figure 1.**
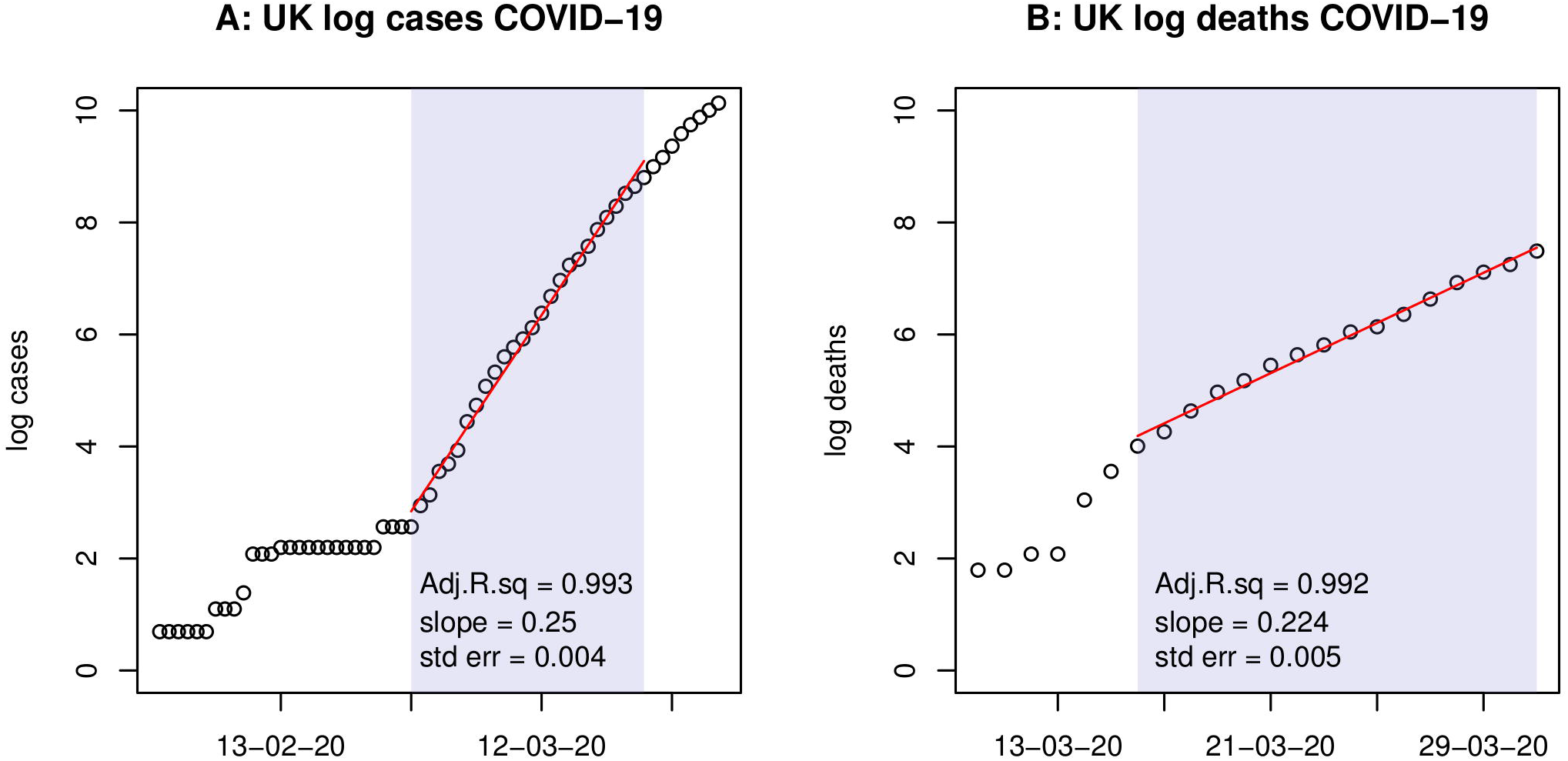
Figure 1 shows the natural logarithms of Public Health England data for (A) cases and (B) deaths up to 31 March. A linear fit to the straight line portion of the log data is shown in red, along with Adjusted *R*^2^ and the estimate and standard error of the slope.

An unpublished implementation of the SEIR model, together with default parameters, clinical evidence, and **R** code is available online from Alison Hill. [8, 9] The code includes functions Getr_SEIR and GetRo_SEIR to compute **r** and **R**_**0**_ respectively, depending on the parameters and population size. The **r** estimate uses the approximation that in the early period of growth, virtually the entire population is susceptible, and the SEIR model reduces to a linear differential equation with growth determined by the largest eigenvalue of its matrix. The model is fitted by numerical integration using the **R** package “deSolve”.

The transmission parameters pertain to patients whose condition is mild, severe, or critical. They are estimated by an adaptive Metropolis-Hastings (M-H) algorithm, fixing all clinical parameters at the values chosen by Hill. A trio of transmission parameters (*b1, b2, b3*) then determines **r** through Getr_SEIR. The likelihood of the trio is taken to be the gaussian likelihood for the resulting value, with mean and sd given by the estimate and standard error of the growth coefficient in M1. A trio receives a higher or lower likelihood if the model gives a fitted log growth rate nearer or further from the empirical value obtained directly from the data. The prior for the trio is set as the product of independent gamma priors for each component, using the same shape and rate for each. Adaptive M-H was run (**R** package “MHadaptive”) to generate a sample of length 50,000 after burn-in and thinning. Convergence was checked by standard diagnostic tests (**R** package “coda”). The output of M-H is a posterior sample of the trio (*b1, b2, b3*). Since **R**_**0**_ is completely determined by the parameters, the rest of which are fixed, the sample generates a sample for **R**_**0**_. The code file paramest1.txt implements these calculations. All code files are available as Supplementary Material.

Forecasting cases and deaths uses the model and parameters, but also requires an initial value for the numbers of Susceptible, Infected, Infectious (mild), Infectious (severe), Infectious (critical), Recovered, Dead, and total Population. As the projections are short term, Population was fixed at N = 66 million. The run was started from 28 February, for which Public Health England shows a cumulative total of 19 cases and no deaths. 3 cases had already been identified by 8 February, so it was assumed here that they had recovered by 28 February. As the UK was only identifying cases in hospital and none had died, the remaining 16 cases were either severe, critical, or recovered. Lacking other information, the remaining 16 cases were assigned as 10 severe, 4 critical, and 2 recovered as of 28 February.

In order to estimate the numbers of Infected and Infectious (mild) on 28 Feb, none of which would have been identified as they were not hospitalised, note that the model assumes all infected cases become infectious (mild). Once the growth rate is estimated for the exponential phase, and given the incubation period, the ratio of Infected to Infectious (mild) is fixed at any time during this phase. Maximum Likelihood estimates were then obtained, subject to that constraint, to minimise the sum of the squares of Pearson residuals for predicted vs observed deaths in the period 28 February – 23 March, the last date before lockdown.

Once the initial values on 28 February are chosen, the model can be run with the transmission parameters set at their mean values from the sample, with reductions imposed during a lockdown period. One scenario, shown in Figure 2, assumed the lockdown would reduce each *b*_*i*_ to 20% of its sample mean. For Figure 3, using the 20% lockdown, 500 samples were drawn for (*b1, b2, b3*). The model was run using each sample to reset the initial conditions and forecast severe and critical cases during the lockdown. Quantiles of the results at each time point then gave 90% credible envelopes. In Figure 4, each *b*_*i*_ was reduced to 10% of its mean during the lockdown.

**Figure 2.**
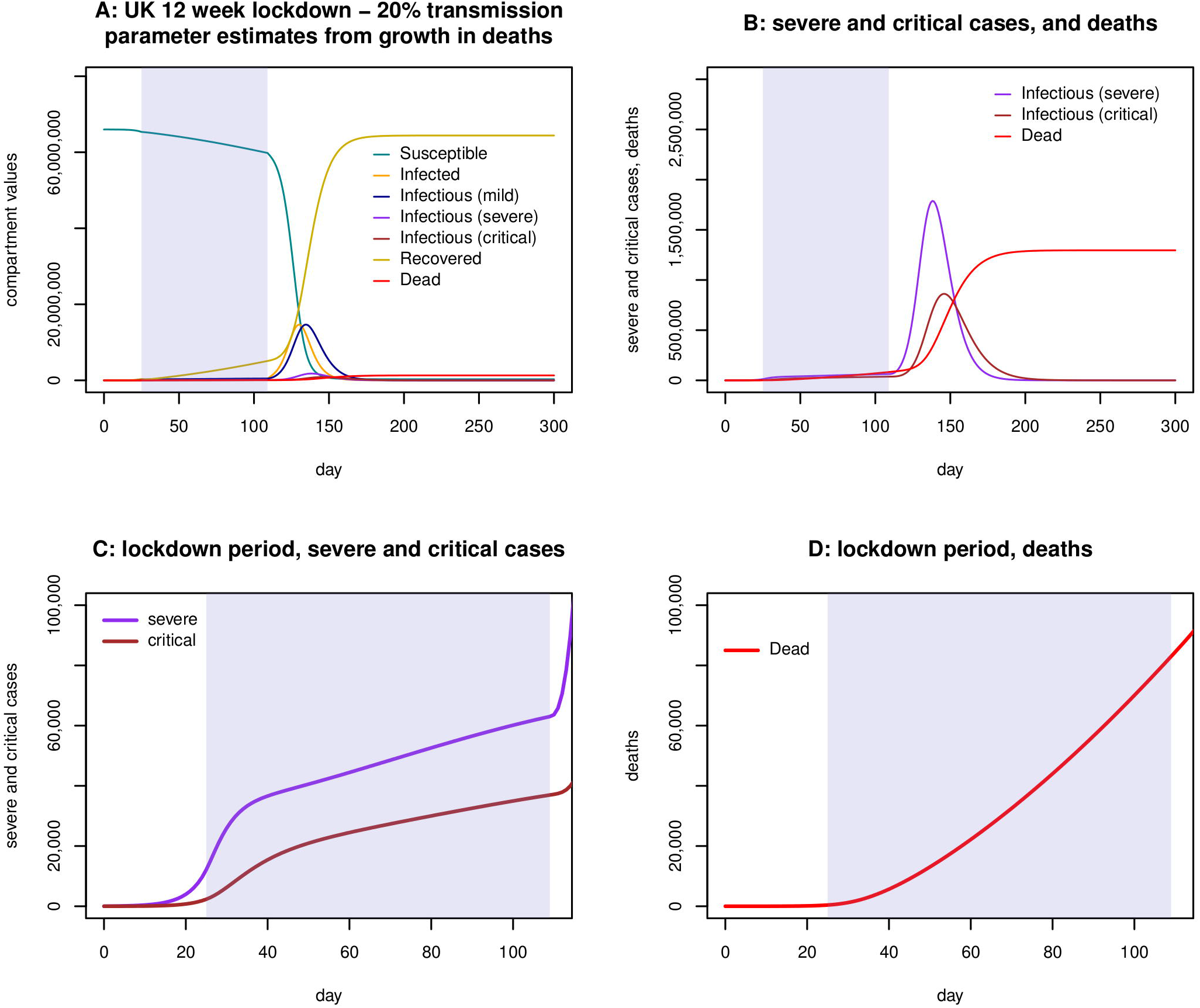
Figure 2 assumes that the lockdown beginning on 24 March cuts all three transmission parameters (*b1*:mild, *b2*:severe, *b3*:critical), to 20% of their previous values, after which they are fully restored. (A) shows the first 300 days of the epidemic, with the lockdown portion shaded. The eruption of cases and deaths is delayed until after the lockdown ends. (B) shows severe and critical cases, and deaths, again for the entire 300 days. (C) shows severe and critical cases rising throughout the lockdown. (D) shows cumulative deaths reaching over 80,000 on 16 June when the lockdown ends.

**Figure 3.**
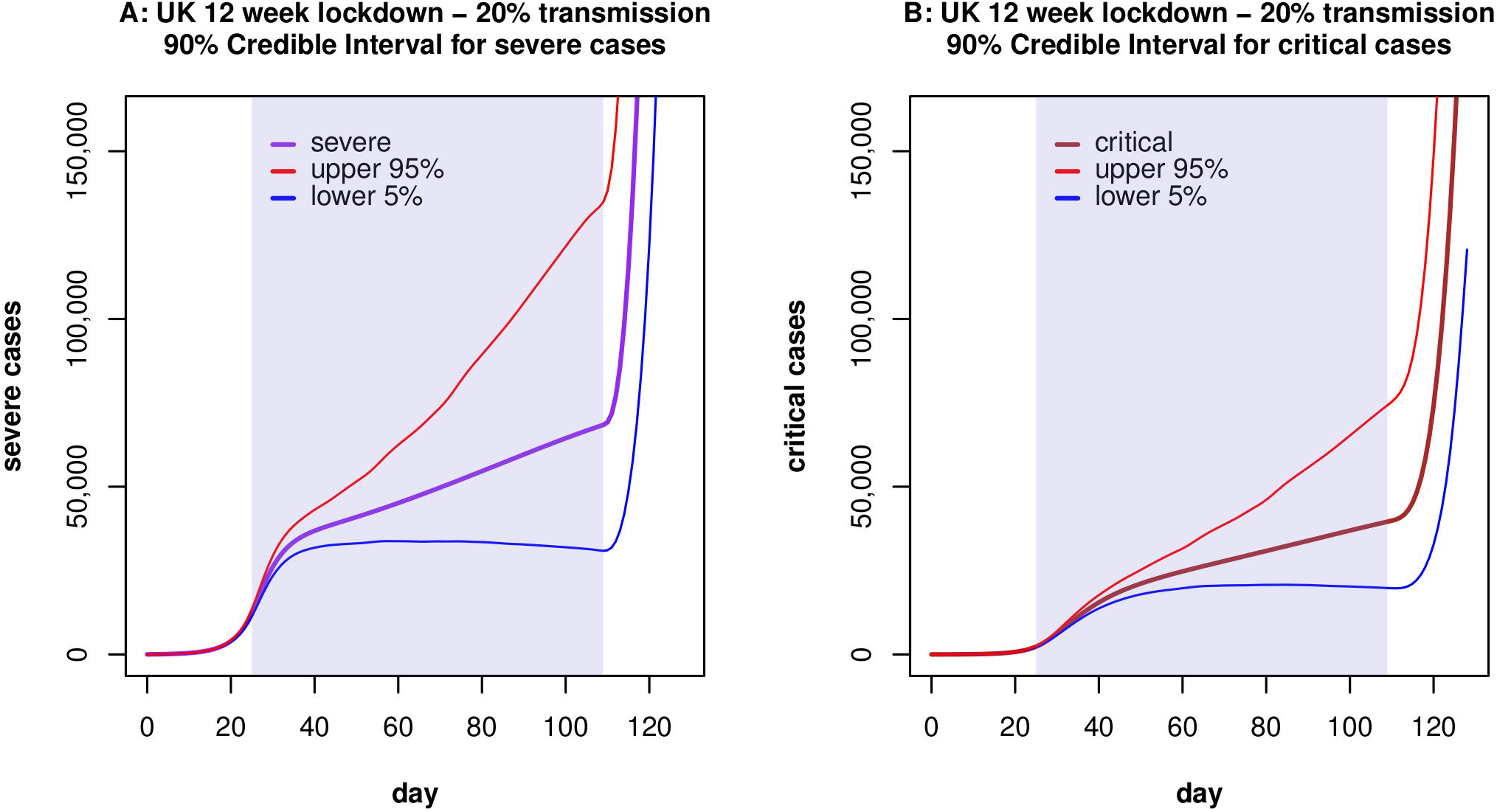
Figure 3 shows 90% credible envelopes for (A) severe, and (B) critical cases during a 12 week lockdown with all transmission parameters reduced to 20% of their previous value. At each time point, the upper 95% and lower 5% quantiles are found using 500 parameter samples from the Metropolis-Hastings algorithm. These indicate the uncertainty arising from transmission parameters whilst ignoring all other variability, whether in the model itself or the clinical parameters.

**Figure 4.**
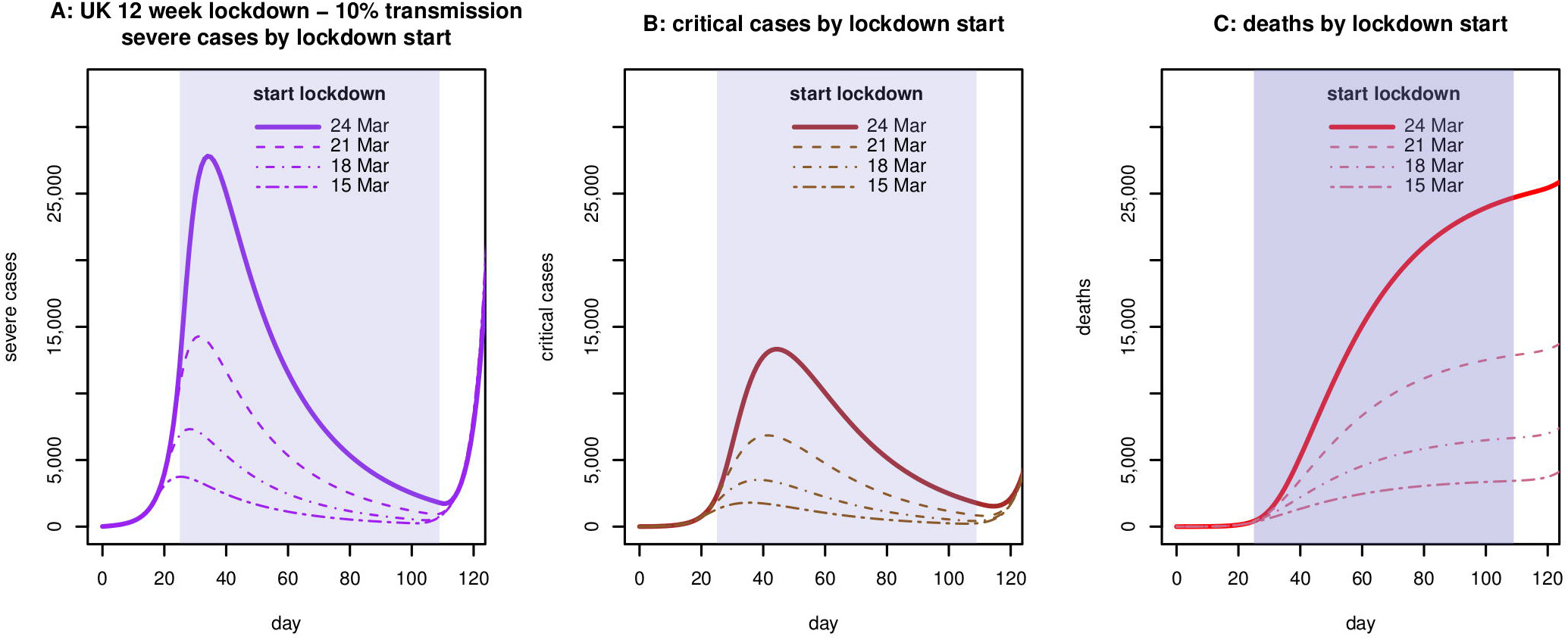
Figure 4 assumes the lockdown will cut each transmission parameter to 10% of its previous value, and compares the forecasts for (A) severe, (B) critical, and (C) deaths using lockdown start dates 24 March (solid), 21 March (dashed), 18 March (dotdash), and 15 March (twodash).

The sensitivity of results was further tested by varying the start date of the 12 week lockdown, or the Case Fatality Rate, or the prior, or the assumed numbers of infected and infectious (mild) cases and assignment of hospitalised severe and critical cases on 28 February.

In the code file paramest1.txt, which generates the parameter samples and estimate and CI for **R**_**0**_, neither Getr_SEIR nor GetRo_–_SEIR depend directly on the parameter u governing the death risk for critical patients, but only on *u + g3* which is the reciprocal of the average length of stay in ICU (see Hill [8]). Thus the estimates for **R**_**0**_ are independent of the Case Fatality Rate. However, the forecasts do depend on CFR.

As this study uses only freely available anonymous data released by Public Health England, ethical approval was not sought.

## Results

From Figure 1, linearity of log(deaths) is clear in the range 16 – 31 March. A linear fit has Adjusted *R*^2^ = 0.992, and the coefficient of growth **r** = 0.224 with s.e. = 0.005. Adaptive M-H sampling passed the Raftery, Geweke, and Heidelberger tests for convergence. Posterior estimates of the transmission parameters, scaled by population N = 66 million, are *b1*\**N* = 0.735 with 90% credible interval (0.576, 0.826) whilst *b2*\**N* = 1.112 (0.055, 3.335) and *b3*\**N* = 1.015 (0.052, 3.071). **R**_**0**_ is estimated as 5.81 with 90%CI (5.08, 6.98).

As the incubation period is taken to be 5 days (see Hill [8]), at any time t during the exponential phase Infectious (mild) [t] = Infected [t-5]. As both are growing with **r** = 0.224, Infectious (mild) [t] = Infected [t] / *e*^5·0.224^ = Infected[t] / 3.06. Applying the estimated parameter values and assigning hospitalised cases on 28 February as 10 severe, 4 critical, and 5 recovered, the constrained Maximum Likelihood estimates for that date are 1627 infected and 532 mild (see Methods) and the predictions give a reasonable approximation to deaths in the period to 23 March.

Figure 2 shows the results of a model run with these initial conditions, and a lockdown assumed to reduce each *b*_*i*_ to 20% of its estimate throughout a 12 week period 24 March – 16 June. Severe cases rise throughout the lockout, reaching nearly 63,000 by the end. Likewise critical cases rise throughout, reaching nearly 37,000, and total deaths exceed 81,000. Figure 3 shows the 90% confidence envelope for the evolution of severe and critical cases during the lockdown. In contrast, as shown in Figure 4 (bold lines), if each *b*_*i*_ falls to 10% of its estimate throughout, severe cases peak on 2 April at just under 28,000 whilst critical cases peak on 12 April at over 13,000 and deaths during the lockdown are nearly 25,000.

### Sensitivity

These results are very sensitive to the starting date of the lockdown. If the 20% lockdown began one week earlier, on 17 March, cases and deaths would still rise throughout the 12 weeks but the totals on 9 June would be around 16,500 severe and around 9,250 critical cases, and 18,500 deaths. For a 10% lockdown beginning on 17 March, severe cases peak on 26 March at under 5,900 whilst critical cases peak on 5 April at 2,800 and total deaths by 9 June are 5,200. Figure 4 includes curves for severe and critical cases, and deaths, if a 10% lockdown had begun 3, 6, or 9 days before 24 March.

The **R**_**0**_ estimate and CI are independent of the Case Fatality Rate (see Methods). The results above and Figures were generated with CFR = 2%, the value shown by Hill [8]. If CFR = 1%, a 20% lockdown beginning 24 March would again result in severe and critical cases rising throughout the 12 weeks, but the totals on 16 June would be nearly 97,000 severe and 60,000 critical with 75,000 deaths. This forecast includes the recalculation of the initial condition on 28 February with the lower value of CFR requiring more infected and infectious (mild) cases to fit the known data on deaths. For a 10% lockdown with CFR=1% severe cases peak on 2 April at over 55,000 and critical cases peak on 12 April at around 26,500 with over 24,000 deaths by 16 June.

**R**_**0**_ depends on the prior, but not excessively. With prior shape = 0.3 and rate = 0.3, **R**_**0**_ is estimated as 6.73 with 90%CI (5.63, 9.63).

The forecasts barely depend on how the 19 known cases on 28 February are assigned, but do depend on the unknown numbers that day of infected and infectious (mild) cases, inferred here from the growth rate in the exponential phase and the numbers of reported deaths. For example, if the ratio of infected / infection(mild) remains at 3.06 but the number of infected cases on 28 February is taken as 2000 rather than 1627, a 20% lockdown from 24 March to 16 June shows severe cases rising to over 72,000 whilst critical cases rise to nearly 43,000 and deaths total nearly 98,000.

## Discussion

This brief analysis uses an established deterministic SEIR model [8] for the development over time in the expected numbers of susceptible, infected, infectious, recovered cases and deaths, depending on parameters, some of which can be estimated from clinical studies since the outbreak of COVID-19. Other parameters concern the rate at which persons who have become infectious will infect others, depending on whether their clinical condition is mild, severe, or critical. Clinical parameters, such as the delay between “infected” and “infectious”, may not vary greatly between countries. By contrast, the rate at which infection is transmitted in the community depends on level and types of social interaction, which may vary over time in response to public policy, such as a lockdown, or weather and season. For hospitalised patients, transmission also depends on the level of protection for healthworkers and environmental controls including cleaning and air quality. If the evolution of all these parameters were known, the model would predict the numbers of people in different stages of the disease, or death, over time. Of course the model itself may be inadequate, no matter how the parameters are chosen. The SEIR model is much simpler than the hierarchical model being developed in the recent report from Imperial College. [10].

In the context of the SEIR model, in the early period of an epidemic, numbers of infected or infectious persons or deaths are all growing exponentially at the same rate, and so the slope of their logs is identical. Given the uncertainty regarding true numbers of COVID-19 cases due to the lack of testing, the analysis here uses Public Health England data on deaths for the period before the lockdown began on 24 March. Even so, the data excludes deaths in care homes. However, if the proportion of hospital deaths as a fraction of all COVID-19 deaths remained constant during the early phase, log(deaths) would simply be offset by a constant, and therefore the estimate and standard error of its slope would be unaffected.

I estimated transmission parameters in the UK, on the assumption that clinical parameters are fixed at the values already estimated by Hill. A likelihood was assigned to transmission parameters and samples obtained via the Metropolis-Hastings algorithm. These samples lead to an estimate and credible interval for the basic reproduction number **R**_**0**_. In reality, the clinical parameters are not fixed and their estimates will develop with new research. Therefore, the samples may be biased by the assumed clinical values and may underestimate the variability of the transmission parameters, so the true CI for **R**_**0**_ in the UK may be wider than reported here.

The value found here is compatible with a recent analysis of global data by Steven Sanche and co-workers (preprint [3]), who estimated **R**_**0**_ in the range 4.7 – 6.6, significantly higher than the value of 3.11 cited by the UK government. [11, 1] **R**_**0**_ itself is based on an idealised notion of perfect mixing, and the analysis here is pooled over the entire population, without stratification by age or any other characteristics.

The parameter samples also enable forecasts, which are not based on **R**_**0**_ but directly on the model and parameter estimates. These forecasts do not include the possibility that asymptomatic patients are infective, and make a crude assumption that transmission was unaffected until the lockdown began, and was then reduced immediately. The model also assumes that length of stay in each compartment is exponentially distributed, but a recent preprint by Robert Verity and co-workers [12] fitted gamma distributions to length of stay, and estimated the coefficient of variation at 0.33 which implies shape=9, rather than shape=1 (exponential). I have not adjusted the model to allow for this.

Whilst the transmission parameter *b1* (mild) influences the spread in the wider community, *b2* (severe) and *b3* (critical) are key to the risk to healthworkers. The estimates for *b2* and *b3* are large, although with wide CIs. Around 80% of cases are mild, and **R**_**0**_ is much more sensitive to *b1* than to *b2* or *b3*, which narrows the CI for **R**_**0**_.

The first lockdown forecast involves an assumption that *b1, b2*, and *b3* each reduce 5-fold, to 20% of their pre-lockdown values. The predicted curves for severe and critical cases rise throughout the lockdown and are set to overwhelm the NHS, where ICU capacity is far below the 20,000 predicted critical cases in mid-April (Figure 3), let alone nearly 37,000 by the end of the lockdown. With transmission reduced 10-fold (Figure 4), severe and critical cases each peak in April, but the pressures will still be intense.

Once the lockdown is lifted, if no vaccine or therapy is yet available and transmission rebounds to its original levels the problem simply recurs as shown in Figure 2 (B). By then the NHS may have more resources.

The delay in beginning a hypothetical 12 week lockdown has a strong effect on the outcome. If the first lockdown scenario began on 17 March rather than 24 March, deaths by the end of 12 weeks would fall from over 81,000 to under 19,000. This is due to the continuing increase of cases during the pre-lockdown period, and raises an unanswered question: why did the UK lockdown only start on 24 March?

## Data Availability

UK data on COVID-19 cases and deaths is freely available from Public Health England

https://fingertips.phe.org.uk/documents/Historic%20COVID-19%20Dashboard%20Data.xlsx

## Acknowledgements

Thanks to Wendy Olsen for helpful comments on drafts, and to Alison Hill for her website.

## Author Contributions

GD conceived of the study and carried out the analysis.

